# Using Mobility Data to Understand and Forecast COVID19 Dynamics

**DOI:** 10.1101/2020.12.13.20248129

**Authors:** Lijing Wang, Xue Ben, Aniruddha Adiga, Adam Sadilek, Ashish Tendulkar, Srinivasan Venkatramanan, Anil Vullikanti, Gaurav Aggarwal, Alok Talekar, Jiangzhuo Chen, Bryan Lewis, Samarth Swarup, Amol Kapoor, Milind Tambe, Madhav Marathe

## Abstract

Disease dynamics, human mobility, and public policies co-evolve during a pandemic such as COVID-19. Understanding dynamic human mobility changes and spatial interaction patterns are crucial for understanding and forecasting COVID-19 dynamics. We introduce a novel graph-based neural network(GNN) to incorporate global aggregated mobility flows for a better understanding of the impact of human mobility on COVID-19 dynamics as well as better forecasting of disease dynamics. We propose a recurrent message passing graph neural network that embeds spatio-temporal disease dynamics and human mobility dynamics for daily state-level new confirmed cases forecasting. This work represents one of the early papers on the use of GNNs to forecast COVID-19 incidence dynamics and our methods are competitive to existing methods. We show that the spatial and temporal dynamic mobility graph leveraged by the graph neural network enables better long-term forecasting performance compared to baselines.

## 1 Introduction

The COVID-19 pandemic has affected almost every country in the world and has resulted in an unprecedented response by governments across the world to control its spread. The social distancing measures are one of the most effective non-pharmaceutical interventions at this stage. The social distancing measures have led to significant change in human mobility behaviour while the mobility change has also affected the disease dynamics inevitably. To better understand COVID-19 dynamics and help to control the disease spread, it is crucial and challenging to provide accurate and timely spatio-temporal forecasting of epidemic dynamics. As machine learning and artificial intelligence(AI) has been successful in many domains, there is an urge to investigate how we can leverage AI-based technologies for infectious disease understanding, modeling, forecasting, and controlling. In this work, we focus on applying AI-based techniques to solve the above challenge by incorporating a new large-scale aggregated spatio-temporal mobility data into graph-based neural networks. Using aggregate mobility data to understand COVID-19 dynamics has received wide interests recently. There have been a number of recent studies along these lines, for example, in China using Baidu data [Chinazzi *et al*., 2020], in the US using mobility data [Kraemer *et al*., 2020; Adiga *et al*., 2020b], and at a global scale using airline traffic [Adiga *et al*., 2020a]. On the other side, lots of COVID-19 forecasting methods are proposed since the initial outbreak early this year, such as mechanistic methods [Yang *et* al., 2020; Anastassopoulou *et al*., 2020; Kai *et al*., 2020], and time series methods using statistical regression models [Ribeiro *et al*., 2020] or deep learning model [Ramchandani *et al*., 2020]. However few of the existing works investigate spatio-temporal forecasting using graph neural network (GNN) with integrated real-time mobility data. In our work, we introduce a novel method to incorporate global population mobility flows into graph-based spatial-temporal neural networks for COVID-19 dynamic forecasting. Our major contributions are:

- We analyze the joint effects of social-distancing guidelines and mobility patterns at US state levels using an integrated map of mobility flows (MF), COVID-19 Surveillance data and data on social distancing guidelines;
- We design a dynamic mobility informed GNN that considers both temporal dynamics and cross-location coevolution dynamics using a recurrent message passing (RMP) module to recurrently embed information from a node’s neighbors;
- We also design multiple variants of the proposed model which use static mobility graph, geographical adjacency graph, and attention-based trainable graph;
- We evaluate the proposed model on forecasting the US state level daily new cases, and demonstrate that the dynamic spatial and temporal mobility informed GNN allows for better forecasting performance compared with its variants as well as several existing classic and state-of-the-art time series methods.

## 2 Methods

### 2.1 Problem formulation

Assuming we have *N* regions in total. Each region is associated with time series of multiple observed features, e.g. surveillance cases, in a time window *T*, where *T* is the observation duration. It could be of weekly or daily granularity depending on the data resolution. We define a dynamic graph of *N* regions as *G*(𝒱, ε, 𝒯), where 𝒱 is the set of *N* nodes, ε ⊆ 𝒱 × 𝒱 is the set of edges, and 𝒯 is the set of *T* time points. A node *v*_*i*_ at time *t* is attributed with 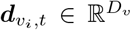 where *D*_*v*_ is the node feature numbers. An edge *e*_*ij*_ ∈ *ε* at time *t* connecting nodes *v*_*i*_ and *v*_*j*_ is weighted by either adjacency matrix or mobility flow matrix, is attributed with 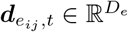 where *D*_*e*_ is the edge feature numbers.

### 2.2 Mobility informed graph neural networks

#### Constructing the graph

We construct a dynamic mobility graph *G*(𝒱, ε, 𝒯), where each node feature ***d***_*v*_ includes a sequence of dynamic observations regarding the the region in a history window *H* ≤ *T*. We include daily new case count, new death count and intra-region mobility flow (*f*_*ii*_ (*t*) which represents the MF from *v*_*i*_ to *v*_*i*_ during time *t*) as the node features, i.e. 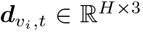. The graph edge features are derived from the inter-region mobility by aggregating Google mobility data to the state or county level. At a certain time point *t*, if there is any human movement from region *j* to region *i* in the past *H* days, we add a directed edge *e*_*ji*_ that connects region *j* and *i*, and associate it with the inter-region mobility flow *f*_*ji*_(*t*) and flow of active cases from source region (defined as 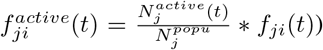 as the edge feature i.e. 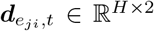 where 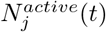 is the number of active cases (cumulative cases minus recovered cases and deaths) and 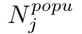 is the population of region *j*.

#### Feature encoding

In the graph, node feature vectors and edge feature vectors include temporal information from the past. We encode the vectors using a Long Short-Term Memory (LSTM) module. At time step *t*∈ 𝒯, for each feature vector 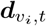 or 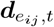 the LSTM module encodes the vector into hidden representations as:

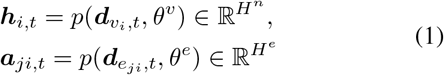

where *p* denotes LSTM cell computation, *H*^*n*^ and *H*^*e*^ are hidden dimension of node feature and edge feature, *θ*^*v*^ and *θ*^*e*^ are parameters to be learned.

#### Spatio-temporal message passing

A region’s COVID-19 dynamics can potentially be affected by regions where frequent travels occur between them. This resembles the core insight behind graph neural network models, i.e. the transformation of the input node’s signal can be coupled with the propagation of information from a node’s neighbors in order to better inform the future hidden state of the original input. This is most evident in the unified message-passing framework proposed by [Gilmer *et al*., 2017]. In our model, we design an *Recurrent Message Passing* (RMP) module to recurrently pass the hidden representations from a node’s neighbors to the current node. As shown in Figure 1, the RMP module has two phases: the message passing (MP) phase and the update phase (UP). It runs for *L* rounds, so any node in the graph is taking into account of neighbors that are *L* hops away. To be more specific, at time *t*, given a node *v*_*i*_ at a certain round (*l* + 1): In the MP phase, for each node pair (*v*_*i*_, *v*_*j*_) that *v*_*j*_ ∈ 𝒩 (*i*) where 𝒩 *i* denotes neighbors of *v*_*i*_, we first combine the node hidden states ***h***_*i,t*_, ***h***_*j,t*_ and in-edge hidden representations ***a***_*ji,t*_ from previous round *l* using a message passing function *f* to get a hidden state ***a***_*ji,t*_ at current round *l* + 1. It will later be aggregated (we use mean operation but can be sum, max, etc.) together over all pairs to obtain a message ***m***_*i*_ for node *v*_*i*_. In UP phase, we use a node upate function *g* to update the the node hidden states. The hidden states of the node *v*_*i*_ at the (*l* + 1)th round 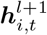 are updated in RMP module as :

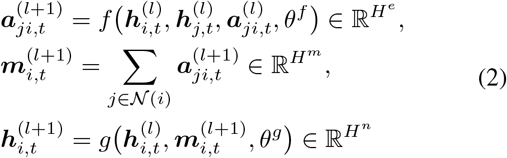

where *θ*^*f*^ and *θ*^*g*^ are module parameters to be learned, 𝒩(*i*) denotes the neighbors of *v*_*i*_ where there exists *e*_*ji*_, *f* is the message passing function that uses a multilayer perceptron (MLP) and *g* is the node update function that uses Gated Recurrent Unit (GRU), ***m***^(*l*+1)^ is the messages passed between nodes.

**Figure 1:**
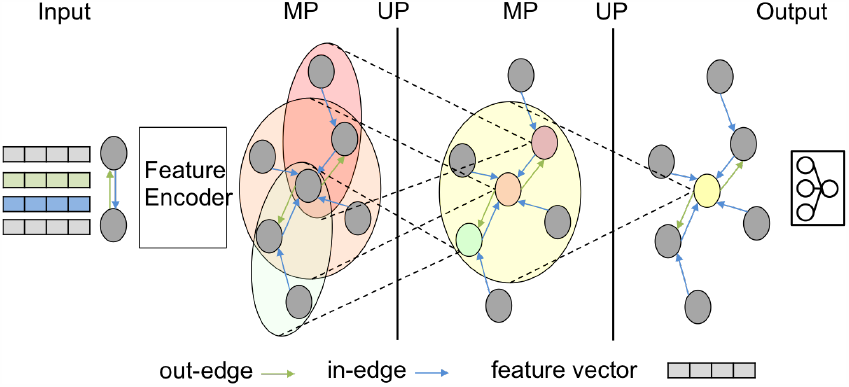
An example of two-hop RMP architecture. Temporal node feature and edge feature vectors are encoded using the feature encoder module. A two-hop RMP module is used to further embed spatio-temporal information to hidden representations. The output module makes the final predictions.

#### Output layer

We feed the hidden representations to the output layer for the final predictions:

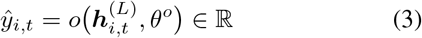

where *θ*^*o*^ is the parameters to be learned, *o* is the output function which is a multilayer perceptron (MLP) in our model.

#### Forward passing process

As shown in Figure 1, we first feed the sequences of temporal node and edge features through the feature encoding module to obtain node and edge embedding, which are utilized as the initial node and edge hidden representations for the RMP module. Then we perform MP and UP computations for *L* rounds. This is the core step to allow one region to leverage information from its neighbors and their connectivity in between. The output module will output the final predictions.

##### Algorithm 1: MF informed GNN forward passing

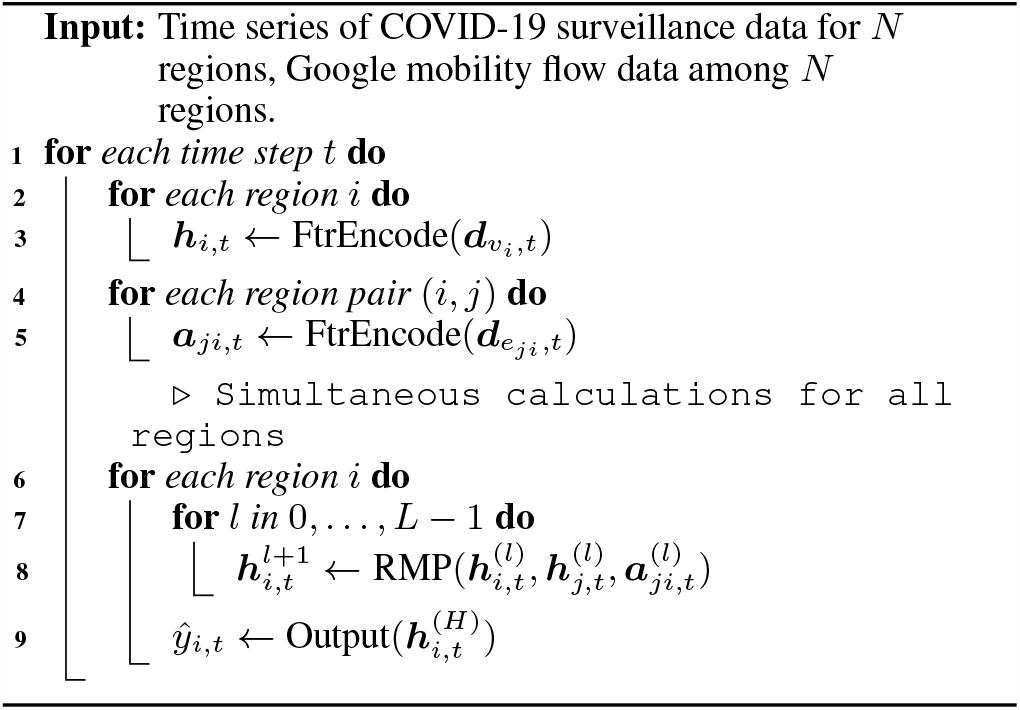

#### Proposed models

The proposed model aims to examine the effect of dynamic mobility on understanding and forecasting COVID-19 dynamics. Thus we design several variants of the proposed model using dynamic mobility graph denoted as GNN-dmob, using a static mobility graph denoted as GNN-smob, using a geographical adjacency graph denoted as GNN-adj, and using an attention-based matrix denoted as GNN-att. The details are listed below.

- **GNN-dmob** the proposed model with dynamic mobility graph.
- **GNN-adj** uses the same graph structure with GNN-dmob but remove intra-region mobility flow from node features i.e. 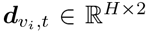, and construct an adjacency matrix by adding an edge *e*_*ij*_ if region *j* is a neighbor of region *i* with edge weight 1. The adjacency matrix is normalized by row summation. It is static across time steps, thus the edge feature is 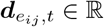.
- **GNN-smob** is similar to the GNN-adj but obtained by replacing the adjacency matrix with a static mobility graph which is an average of mobility graphs from March 1st, 2020 to August 2nd, 2020.
- **GNN-att** is inspired by cola-GNN proposed in [Deng *et al*., 2019]. Instead of using a physical matrix in our model, we implement an attention-based model that allows the model to learn an attention matrix of all the regions. In MP phase, we update the message between nodes as:

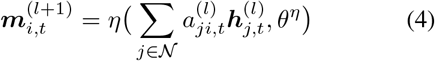

where *θ*^*η*^ is the trainable parameters, *a*_*ji*_ is the attention coefficient defined as: 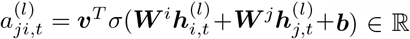 where 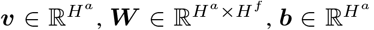 are trainable parameters, *σ* is Rectified Linear Units (ReLU) applied at element-wise.

## 3 Experiments

### 3.1 Datasets

**Google COVID-19 Aggregated Mobility Research Dataset**, which contains the anonymized relative MF aggregated weekly over users within a 5 km^2^ cell. **COVID-19 surveillance data (CSD)** via the UVA COVID-19 surveillance dashboard [UVA, 2020 accessed August 29 2020]. It contains daily confirmed cases and death count worldwide. The data is available at the level of a county and state in the US. Daily case counts and death counts are further aggregated to weekly counts.

### 3.2 Metrics

The metrics used to evaluate the forecasting performance are: *root mean squared error (RMSE)* and *Pearson correlation (PCORR)*.

### 3.3 Baselines

We implemented several classic and state-of-the-art time series models as the comparison methods. They are Naive (uses the observed value of the most recent time point as the future prediction), Autoregressive (AR), Autoregressive Moving Average (ARMA), Long Short-term Memory (LSTM), CNNRNN [Wu *et al*., 2018], and cola-GNN [Deng *et al*., 2019].

### 3.4 Settings and implementation details

The weekly mobility graph is expanded to daily by dividing the weekly values by 7. The training data set is from March 1st to August 1st (125 days), the testing set is from August 2nd to August 29th (28 days). We make 2, 7, 14, 21, and 28 days ahead forecasting for each data point in the testing set. For all models, the historical window *H* = 28. For GNN-dmob, we use a single layer LSTM for feature encoding with 16 units, a two-layer MLP in MP phase with 32 and 16 units, and a single layer GRU in UP with 16 units. The same settings are used for GNN-smob, GNN-att, and GNN-adj. AR and ARMA use AR order 28 and ARMA uses MA order 2. CNNRNN and cola-GNN set with their best parameter settings in the original paper. We set batch size as 32, epoch number as 1000. MSE loss and Adam Optimizer with default settings and early stopping with patience of 100 epochs are used for all model training. All results are average of 5 random runs.

### 3.5 Research findings

In this section, we present key findings and results.

#### Spatio-temporal analysis of MF patterns and COVID-19 dynamics in the US

In order to analyze the human mobility and COVID-19 dynamics during different phases of the pandemic, we use a 4-week window and moving one week ahead each time to compute Pearson correlation between new confirmed cases and MF within the window. Figure 2 shows the Pearson correlation along the weeks. We observe that mobility flow and new confirmed cases show very high negative correlation (median −0.97) for almost all states during March when most of states mandated school closure and stay-at-home orders. This indicates that COIVD-19 dynamics and human mobility are highly correlated. Starting from the mid April when the states started to reopen to some degree, there is a large variation in correlation values, which indicates that COVID-19 dynamics varies a lot due to that it is affected by multiple complicated factors like local population size, individual behaviours (e.g. wearing a mask or not in public location), and government reopening guidelines.

**Figure 2:**
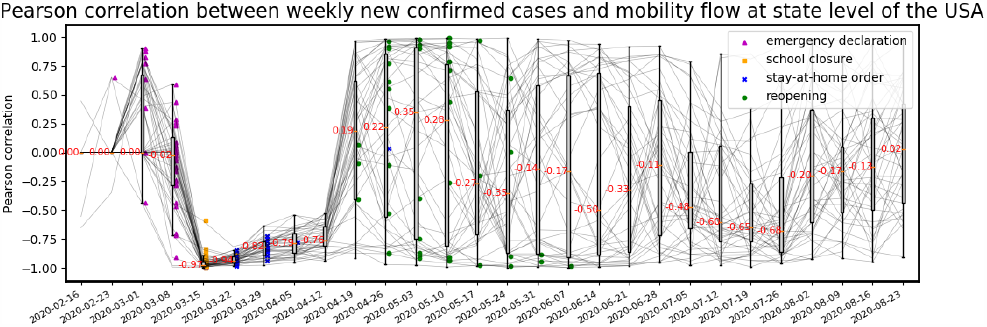
Pearson correlation between MF and new confirmed cases, together with state level social distancing mandates including emergency declaration (purple), school closure (orange), and stay-athome (blue) are marked. A boxplot is used to display variation in samples of 53 states each week. The median value is shown along with the median line.

#### Forecasting performance and discussion

The proposed GNN-dmob model is evaluated w.r.t. forecasting daily new confirmed cases at US state level for 2, 7, 14, 21 and 28 days ahead. Table 1 presents the RMSE, and PCORR performance averaging across 53 states and 28 days. In general, we observe that GNN-dmob has better RMSE performance than the comparisons for long-term forecasting. The best performances on PCORR are evenly distributed among the proposed models. The results indicate that our proposed methods can capture the disease dynamic in both short-term and long-term. Naive baseline outperforms the other baselines for 7, 14, 21, 28 days ahead forecasting. This is not surprising because the testing data is from August during when the time series of all states are showing downward trends with small changing rates. The Naive assumes certain level of regularity in the time series leading to good forecasting performance on the testing data. DNN-based models perform better than AR-based models especially on long-term forecasting which indicates that DNN-based models have better generalization capability for forecasting unseen data. In our experiments, attention-based models cola-GNN and GNN-att are not outstanding for both short and long-term forecasting. A possible reason is that the learned attention coefficients are outdated due to the fast evolution in COVID-19 dynamics between training time period and testing time period (see Figure 3), which leads to false attention by regions while predicting. The proposed method GNN-dmob explicitly projects the most recent mobility patterns to the future potentially leading to better performance.

**Table 1:**
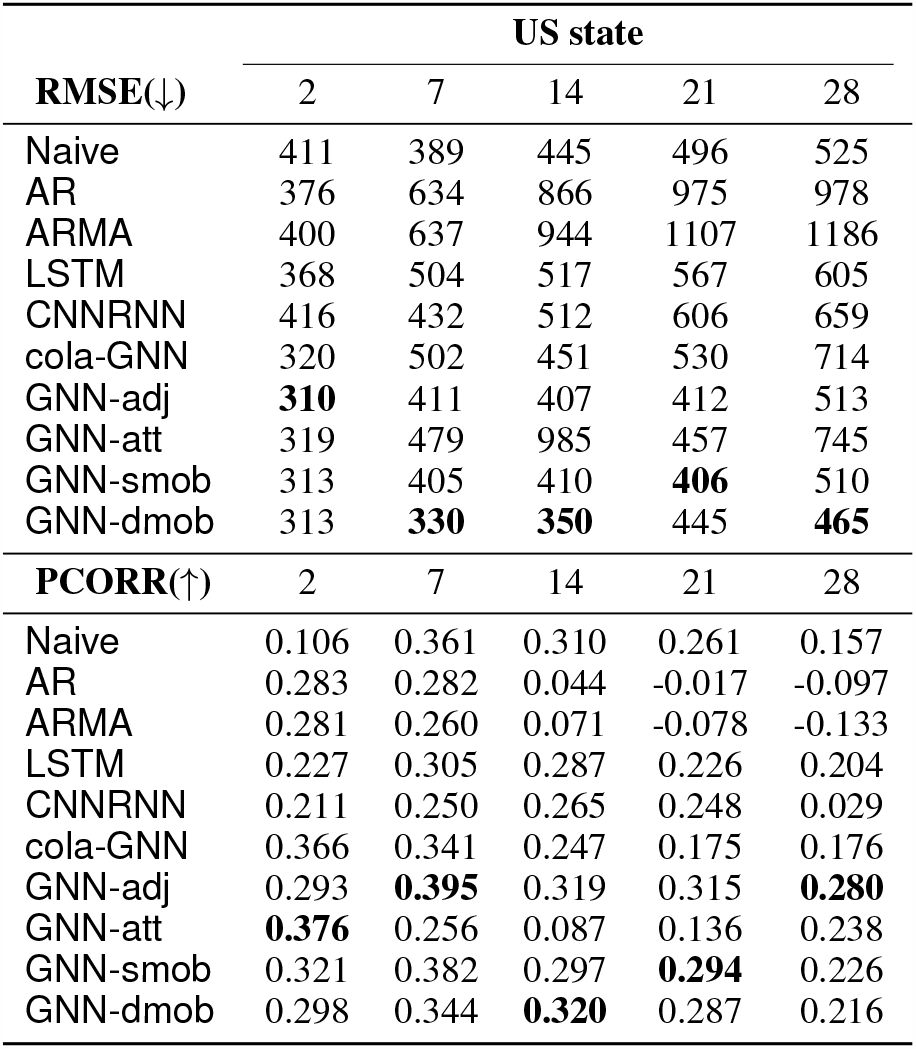
RMSE and PCORR performance of different methods on the US state dataset with horizon = 2, 7, 14, 21, and 28. The values are average of 5 runs. Bold face indicates the best results of each column.

**Figure 3:**
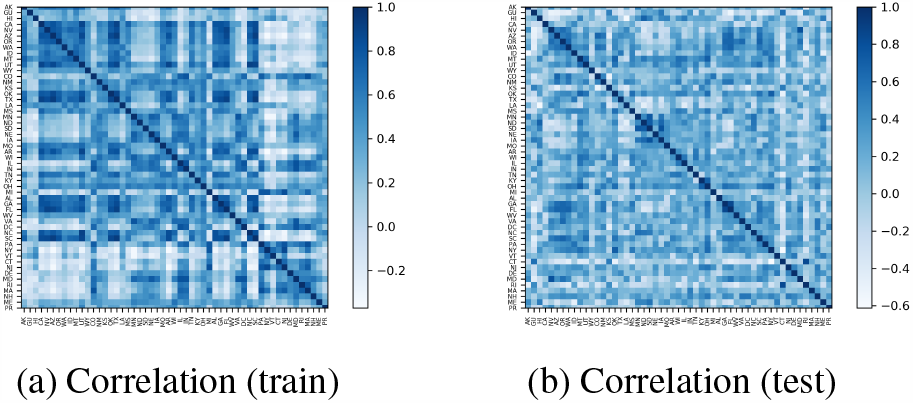
Heatmap of Pearson correlation matrix of state level time series of new confirmed cases. (3a) training data and (3b) testing data. We can observe that the pattern of correlations between states changed dramatically from training dataset to testing dataset.

## 4 Conclusion

The paper introduces a novel GNN framework to incorporate aggregated mobility flows for better understanding the impact of human mobility on COVID-19 dynamics as well as better forecasting of disease dynamics. We propose a recurrent message passing GNN to embed spatio-temporal disease dynamics (COVID-19 surveillance data) and human mobility dynamics (MF data) while making forecasting. The experiment results of forecasting daily COVID-19 new cases for each state in the US demonstrate the additional improvements obtained by using the mobility data. The use of GNN for COVID-19 is just beginning and our results are some of the first results in this area to yield good performance.

## Data Availability

The Google COVID-19 Aggregated Mobility Research Dataset used for this study is available with permission from Google, LLC. University of Virginia COVID-19 Dashboard that provides detailed, global epidemic surveillance data and used in this study is available for public use on \url{https://nssac.bii.virginia.edu/covid-19/dashboard/}. The data on social distancing guidelines in US states and India COVID-19 surveillance data would be shared upon request and would be uploaded on \url{https://dataverse.lib.virginia.edu/}

## Acknowledgments

The authors would like to thank members of the Biocomplexity COVID-19 Response Team and Network Systems Science and Advanced Computing (NSSAC) Division for their thoughtful comments and suggestions related to epidemic modeling and response support. We thank members of the Biocomplexity Institute and Initiative, University of Virginia for useful discussion and suggestions. We also thank Aaron Schneider, Aaron Stein, Ahmed Aktay, Alvin Raj, Amy Chung-Yu Chou, Andrew Oplinger, Ashley Zlatinov, Blaise Aguera y Arcas, Bryant Gipson, Charina Chou, Christopher Pluntke, Damien Desfontaines, Eric Tholome, Ewa Dominowska, Gregor Rothfuss, Iz Conroy, Janel Thamkul, Janet Whiteman, Jason Freidenfelds, Jeff Dean, Karen Lee Smith, Katherine Chou, Leeron Morad, Lizzie Dorfman, Marlo McGriff, Mia Vu, Michael Howell, Paul Eastham, Rishi Bal, Royce Wilson, Ruth Alcantara, Shawn O’Banion, Stephanie Cason, Thomas Roessler, Vivien Hoang and Yanning Zhang for their support and guidance. This work was partially supported by National Institutes of Health (NIH) Grant R01GM109718, NSF BIG DATA Grant IIS-1633028, NSF Grant No.: OAC-1916805, NSF Expeditions in Computing Grant CCF-1918656, CCF-1917819, NSF RAPID CNS-2028004, NSF RAPID OAC-2027541, US Centers for Disease Control and Prevention 75D30119C05935, DTRA subcontract/ARA S-D00189-15-TO-01-UVA. Any opinions, findings, and conclusions or recommendations expressed in this material are those of the author(s) and do not necessarily reflect the views of the funding agencies.

